# “Dynamic Analysis of Heart Rate Variability through Motion Path Analysis of the Poincaré Plot”

**DOI:** 10.1101/2025.03.21.25324311

**Authors:** Madhumanti Thakur, Damodar Prasad Goswami, Kaushik Sarkar, Arnab Sengupta

**Author notes:** Corresponding Author: Prof. Arnab Sengupta, DEPT. OF PHYSIOLOGY INSTITUTE OF POSTGRADUATE MEDICAL EDUCATION & RESEARCH. 244 AJC BOSE ROAD. CALCUTTA *–* 700020. INDIA. Sources of support in the form of grants: None. Conflict of interest: Nil.

## Abstract

Autonomic Nervous System (ANS) maintains body’s homeostasis by regulating heart rate and other vital functions. Heart Rate Variability (HRV) analysis is a non-invasive method used to assess ANS activity by measuring variations between consecutive heartbeats.

Poincaré plot, a widely used HRV analysis tool, visualizes the relationship between successive heartbeats. However, conventional Poincaré plot analysis primarily relies on static statistical descriptors, such as SD1 and SD2, which measure short-term and long-term variability but fail to capture the dynamic progression of heartbeat intervals.

This paper introduces motion path analysis of the Poincaré plot, a novel approach that tracks how heartbeat intervals evolve over time. By following the sequential movement of data points, this method reveals both short-term and long-term HRV trends.

Our study demonstrates that motion path analysis detects subtle changes in heart rate patterns under different physiological conditions, such as rest and stress (e.g., head-up tilt tests). This technique offers a more comprehensive understanding of ANS activity by visualizing point-to-point transitions influenced by sympathetic and parasympathetic responses.

Motion path analysis offers a new way to visualize and interpret the complex temporal fluctuations of ANS and has potential applications in clinical research, providing deeper insight into autonomic regulation in health and disease.

## Introduction

The Autonomic Nervous System maintains an intricate stability of the involuntary physiological processes of the body including heart rate and blood pressure. The autonomic integrating center is located in the central nervous system, where afferent signals from different organs of the body, both voluntary and involuntary, converge and fulfill the ultimate goal of maintaining homeostasis.

It is therefore, comprehensible, that Autonomic Nervous System related diseases will lead to disruption of homeostasis. Evidence shows that various parameters of the autonomic output pattern can predict ANS dysfunction even before their clinical onset. [1].

So, analysis of ANS profile can predict risk factors for ANS diseases [2]. It is, at the same time, useful for detection, evaluation of prognosis and an indication of therapeutic feedback. [3].

The usual battery of tests used to analyze the ANS profile [4] have some inherent constraints. ANS acts on the effector organs in an uninterrupted fashion and is constantly altering, whilst also maintaining homeostasis. All the standard tests for assessment of ANS profile are expressed through a single, isolated numerical value. Hence, the dynamicity of the ANS output pattern remains undetected and unexplored.

One of the tests for estimation of ANS function is examination of cardiovascular autonomic reflexes [5]. The fact that the ANS output pattern is always changing, is clearly detected in the delicate but active variation of heart rate.

There is beat-to-beat variation in heart rate during normal sinus rhythm, and this variation is again altered in healthy and diseased states. Several physiological entities interact continually in order to regulate the instantaneous heart rate. Assessment of this variation in heart rate may indicate some ongoing diseases or may be indicative of some forthcoming disease. As a consequence, heart rate variation analysis has now become a favored tool for assessment of autonomic nervous system functioning. [6].

Heart Rate Variability (HRV) is defined as the variation of the time period between two successive heartbeats over time. HRV largely depends on extrinsic control of heart rate and accurately denotes the interaction between sympathetic and parasympathetic branches of the ANS [7, 8].

Heart rate regulation can be thought of as a coupled network of oscillators [9]. Scientists have tried to explain the two branches of autonomic nervous system from this point of view. In one of our earlier works, we tried to separate these two branches in a specific scenario [10].

Fluctuation in autonomic activity, as seen in Respiratory sinus arrhythmia or vasomotor oscillation, cause alteration in heart rate [11, 12]. The usual parameters used to quantify HRV fall within the categories of time domain (e.g. SDNN, pNN50, RMSSD etc), frequency domain (e.g. HF, LF, HF/LF etc.) and non-linear domain (e.g. Information entropy analysis, SD1, SD2 of Poincaré plot etc.). Time domain parameters represent the variation in time of a particular signal during the examination interval. Frequency-domain parameters determine the absolute or relative amount of signal energy within constituent bands and compute the frequency components of the signal. Non-linear measurements quantify the random and nonlinear behaviors of the RR interval time series [13].

However, expression of various objective parameters of autonomic profile in a numerical form seems to be inadequate. This is because the ANS exerts characteristically its function on the effecter organ in a tonic and continuous manner. ANS output pattern is a tonically active and rapid physiologic parameter. It is delicately tuned with the finer alteration of homeostatic state and is a rapidly changing dynamic homeostatic parameter. So, a single numerical entity of autonomic profile fails to reflect the ever-changing temporal dynamicity of ANS output pattern. It just provides a snapshot of the situation in static mode. In the current paper, we intend to propose a novel descriptive and visual way of expression of autonomic tone that can adequately capture the instantaneous beat-to-beat as well as long term temporal variation of ANS.

**Poincaré plot**, named after Henri Poincaré, is a type of recurrence plot used to visualize periodic or near-periodic nature of a dynamical system. It is also known as a return map that represents a Plot of a time series as a function of the current and previous values.

Given the time series i(*n*), *n =* 1, 2, … N, a 2-dimensional return map is obtained by plotting the time series in a space where the x-axis is i(*n*) and the y-axis is i(*n*+1).

If i(*n*) is the output of a dynamical system and the observation time N is sufficiently long, the characteristics of the phase space of the system can be visualized in the return map.

Poincaré Plot is a point cloud constructed from a set of points and in the two dimensions, the x-coordinate of the point is obtained from a given one-dimensional sequence of numbers and the y-coordinate is the immediate next number. However, instead of taking the immediate next, we can consider a distant point also for lagged Poincaré Plots. The distance is expressed through the lag. Similarly, for a three-dimensional Poincaré plot, we need to generate three dimensional points from the given one-dimensional sequence. This plot was first introduced in the field of environmental science [15] and later applied to electrophysiology [16–19] to evaluate the temporal behavior of heart.

The shape of the cloud of points is generally in the shape of Comet, Torpedo, Fan etc. and those shapes can be visually recognized and interpreted in the context of physical events, depending on their visual form and correlated with various patho-physiological conditions [20]. We can also extract various parameters from this plot depending on our necessity and thus quantify it to have a better understanding.

If an R-R interval becomes longer than the preceding one, it will represent a point above the line of identity in the scatter plot. Similarly, a point below the line of identity indicates the reverse. From another perspective, dispersion of the points along the line of identity reflects long-term variability whereas dispersion perpendicular to the line of identity reflects short-term variability. The delay used to compute the plot has a significant role to look at the system as a dynamical system [21,22]. In fact, the plot with ‘delay one’ in ‘dimension two’ represents the typical Poincaré plot [21–23] which is used to depict short and long term variability [23–30]. The special advantage of this plot lies in its ability to identify beat-to-beat cycles and patterns present in a data series that are very difficult to deal with conventional time and frequency domain analysis [31,32].

Each point of the Poincaré cloud represents a distinct state of the tonic pattern of Autonomic Nervous System, having a specific combination of sympathetic and parasympathetic attributes. The movement from one point to another indicates a shift of this combination status and during normal resting state this is happening continuously. Accordingly, each point of the Poincaré plot can be considered as a transition state in a complex dynamical system, and the pattern of transition from one state to other may express some crucial insight in understanding the spatio-temporal dynamics of the Autonomic Nervous System in health and in disease.

Also, the dynamics of fluctuations in physiological rhythms that exhibit long-term correlation and memory [32], can also be assessed by incorporating various degrees of ‘lag’ in these recurrence plots.

### Standard Quantitative Descriptors of Poincaré Plot

The standard descriptors of the Poincaré plot based upon the ellipse-fitting protocol are shown in Fig. 1. The major axis of the fitted ellipse is aligned with the line of identity, i.e. the line passing through the origin with slope 45*^◦^*, and the minor axis is perpendicular to the line of identity which has a slope of 135*^◦^*and passes through the centroid of the plot. Hence, the major and minor axes of the fitted ellipse can be expressed as 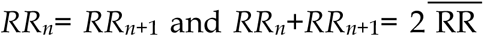 respectively, where RR represents the RR interval series used in the Poincaré plot and 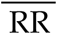 represents the mean value of RR interval series. In the ellipse-fitting protocol, the dispersion of the points along the minor axis measures the width of the plot (SD1), whereas the dispersion of the points along the major axis measures the length of the plot (SD2), and is represented by

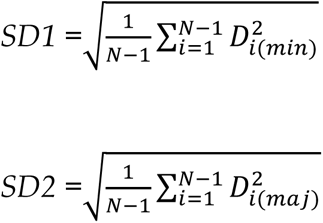

*D*_*i*(*min*)_ *and D*_*i*(*maj*)_ represent the distance of ith. point of the plot *P*(*RR_i_,RR_i_*_+1_), from minor and major axes respectively, and *N* is the number of RR intervals in the taken HRV sequence [33].

**Fig. 1:**
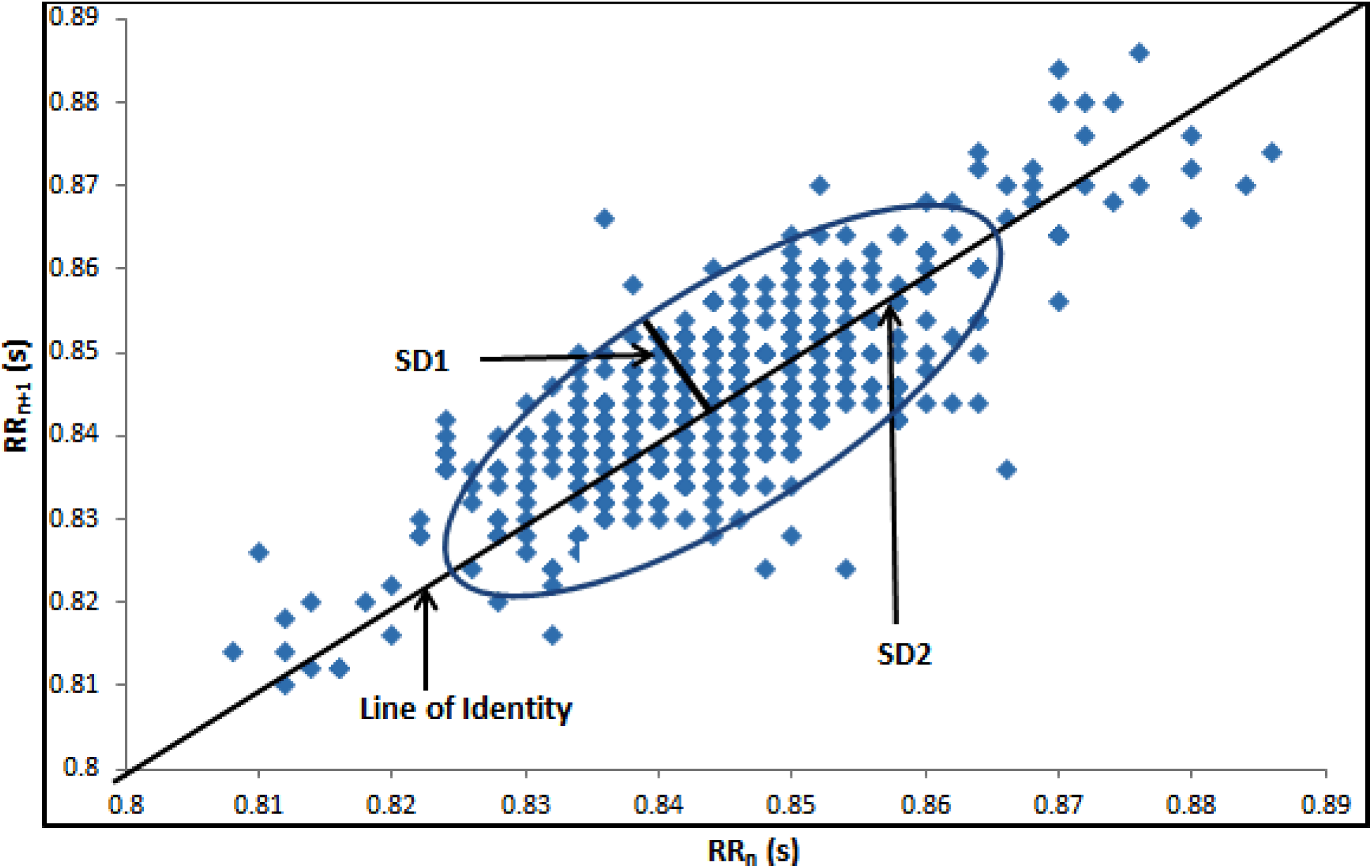
Poincaré Plot.

### Limitation of Standard Descriptors of Poincaré Plot

The derivation of various parameters of the Poincaré plot fundamentally resides on the morphology and distribution of the scatter plot as an image or a stationary entity. The sequence of appearance of the scatter points and their mutual variations with respect to time has not been investigated much, but it may be of substantial relevance to understand the temporal dynamics of the autonomic tone, and capturing temporal variation at point-to-point level of the plot.

It is noteworthy that, as the Poincaré Plot is a phase space representation of the underlying dynamics, the notion of time gets lost there. *SD1* and *SD2* represent the distribution of the points in 2D space and carry only spatial (shape and size) information. It should be noted that many different RR interval series may generate identical plots with exactly similar *SD1* and *SD2* values irrespective of different temporal structures [34], because the shape and size of the plot cannot capture the dynamics of the time series and point to point variation. Fig. 2 represents the pattern of appearance of progressively accumulating scattered points in Poincaré Plot, with successive pair of points of the RR interval time series. This dynamism is clearly expressed in animation form in Video 1(Supplementary material).

**Fig. 2:**
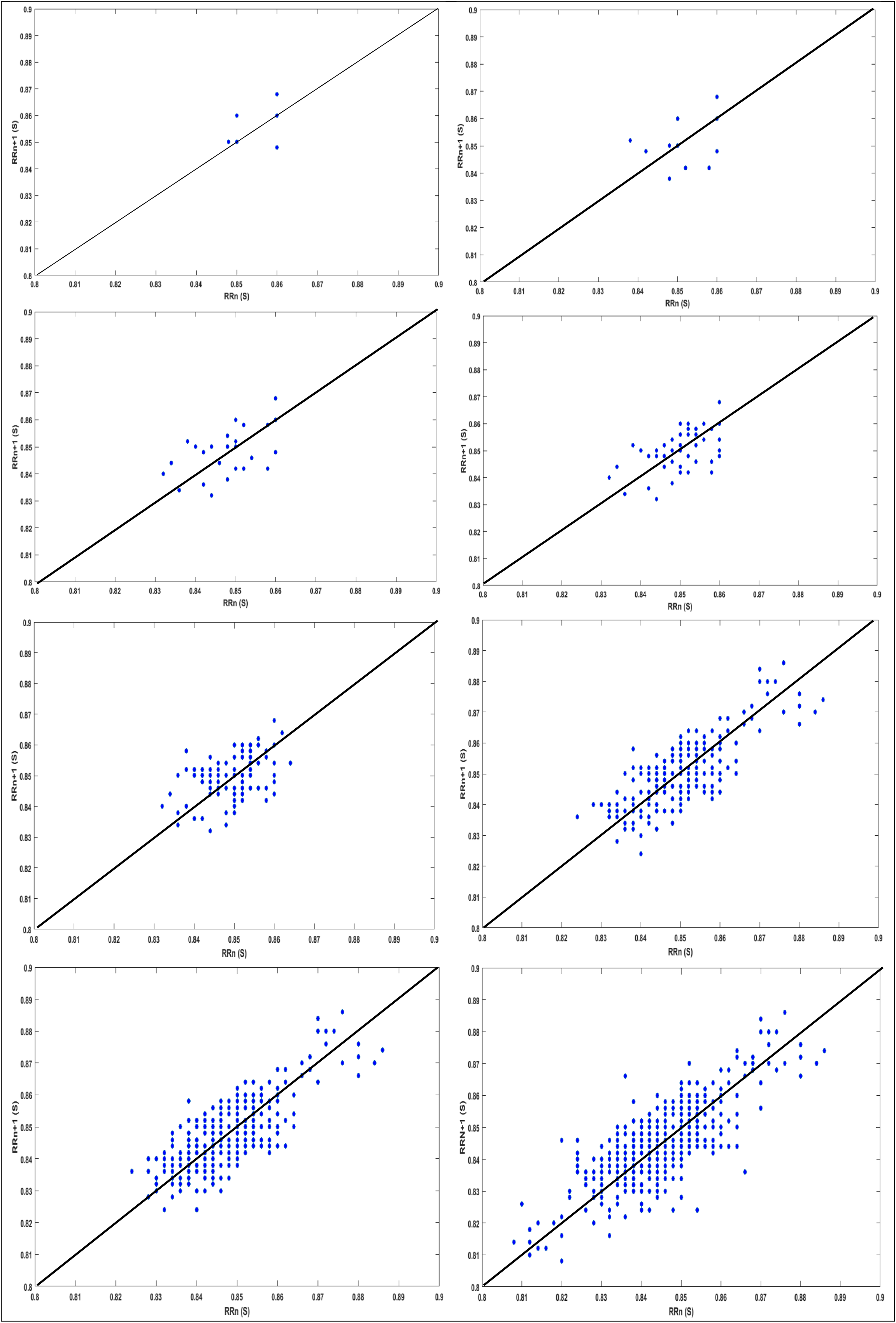
Progressively accumulating Scatter points in Poincaré Plot.

### Motion path Analysis of Poincaré Plot Geometry

One way to study the temporal aspect of the Poincaré Plot is to trace and explore the path traversed by successive scatter points of the plot *in seriatim.* It may be obtained by simple computation and can be plotted as a dynamic attribute of the point-cloud.

In the present article, we propose that, the motion path analysis of the successive points of the Poincaré scatter cloud, as a tool to describe and characterize the temporal dimension of the RR interval time series. At the same time, changes brought about in the temporal dynamics by application of physiological stress on the autonomic tone can also be objectively evaluated.

### Methodology

Motion path evaluation of the Poincaré Plot is carried out in order to analyze the beat-to-beat variation of the RR interval time series data. It is proposed to be conducted in the normal control volunteers without any apparent problems of autonomic nervous system, or any problem or disease state directly or indirectly affecting the autonomic profile.

Poincaré Plot is constructed from the time series data of RR intervals extracted from routine ECG of normal volunteers recorded continuously during rest and also when exposed to orthostatic stress during head-up tilt in a tilt table.

After obtaining the Poincaré plot, the motion path of the successive serial scatter points is reconstructed and attempted to visualize the pattern of movement of point about the line of identity and the animation is represented in .mp4 format [Fig. 3, Video 2 in Supplementary material].

**Fig. 3:**
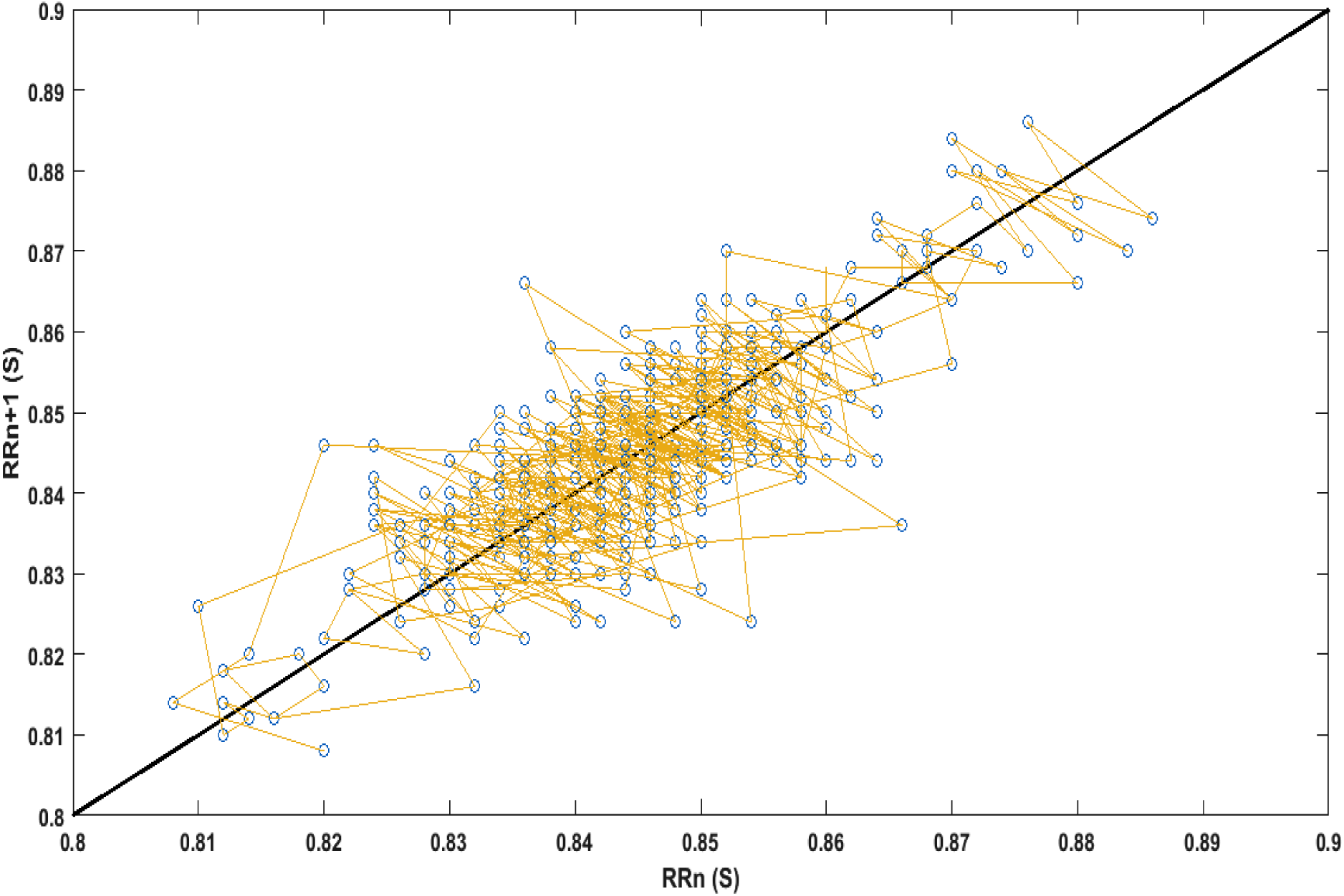
Motion path of the Poincaré Plot.

The dynamic Poincaré plot exhibits a visual movement of the plotted points through the line joining the successive scatter points of the Poincaré plot *in seriatim,* in the form of a motion path. Through this dynamic representation, the extent and pattern of crossing of the line of identity by the cursor provides a crucial insight into the ever-changing dynamicity of the temporal variations of autonomic tone in general and cardiac autonomic activities in particular. At the same time, the overall movement of the scatter points along the longitudinal direction of the Line of Identity represents the pattern of long-term regulation of ANS.

A preliminary finding is illustrated in Figures and Videos (Supplementary material).

Fig. 3 and Video 2 (Supplementary material) depict the Motion path analysis of the Poincaré geometry that visually confirms the phenomenon of beat-to-beat variability of cardiac chronotropy, where movements of the motion path of the progressively accumulating scattered points repeatedly crossing the line of identity from both sides and have an overall spread along the longitudinal axis.

Fig. 4 and Video 3a & 3b (Supplementary material) show the motion path of the progressively accumulating points of the Poincaré plot on application of the slow head-up tilt utilized as orthostatic stress in the present study. It shows that, during the application of stress intervention, the progressively accumulating points of the Poincaré plot form a cluster and gradually move towards the origin of the line of identity.

**Fig. 4:**
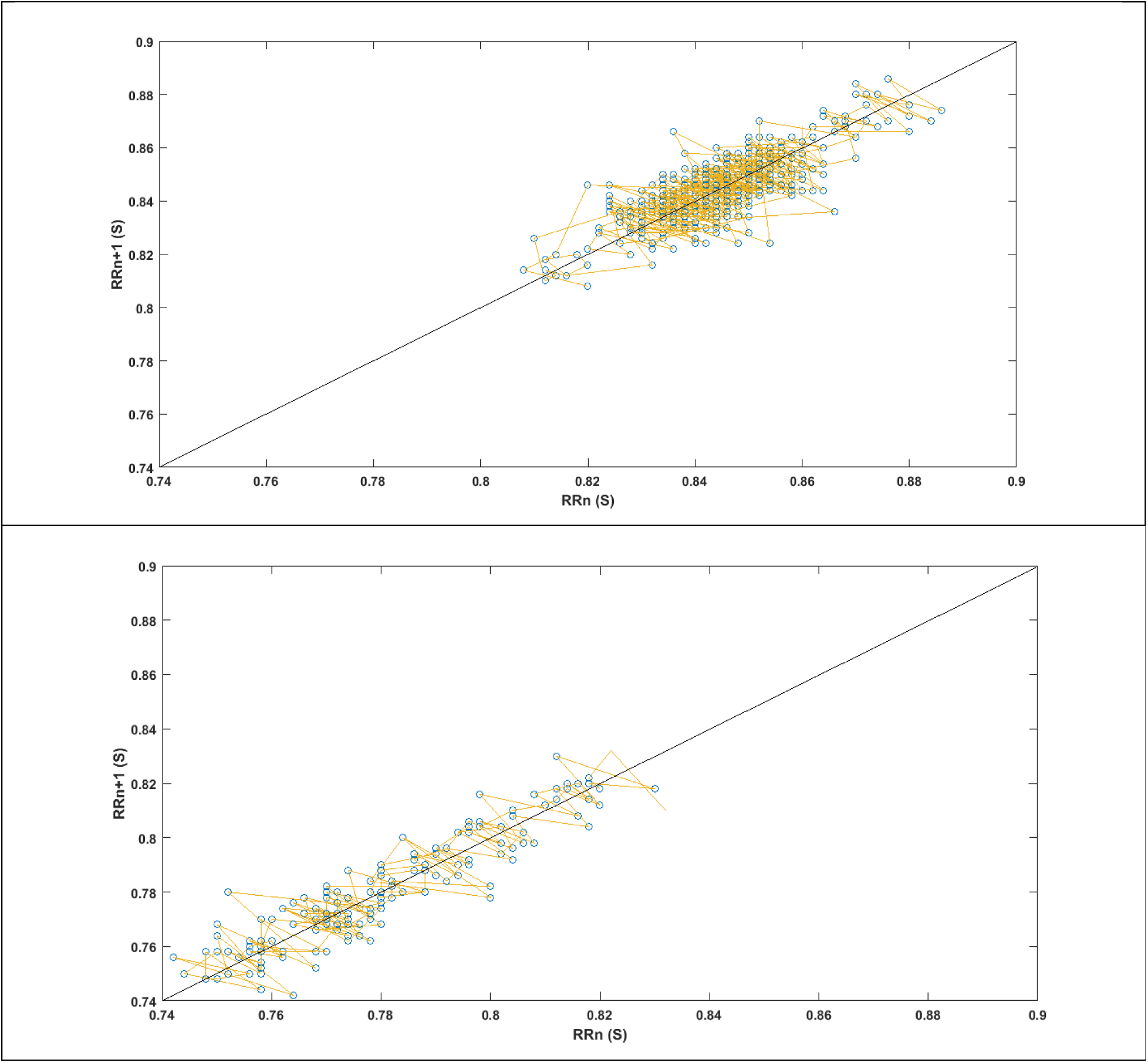
Motion path of the Poincaré Plot during Rest (Upper) & Head-up Tilt (Lower)

It is clearly visible that, the application of the slow orthostatic stress increases the heart rate in a sustained manner that reduces the length of RR intervals gradually. This moves the Poincaré plot cluster towards the origin of the line of identity. It is possible to qualify and quantify this sympathetic effect on ANS tone by analyzing the pattern of motion path accrued.

## Discussion

Recurrence plot is a geometric representation of a time series in the Cartesian plane. Points of the plot are duplets of the values of the time series and the distance between points chosen to form the pair is the *lag* of the plot. Lag-1 recurrence plot, the Poincare Plot displays the autocorrelation of the sequence in a graphical manner. It is a representation of RR interval time series in phase space [35].

It is reasonable that, the cursor of the motion path will move widely in case of large variation of successive RR interval, i.e. in high HRV and the average displacement of the cursor will be smaller in case of low HRV.

The area of the ellipse on the Poincaré plot becomes larger with overall large displacement of the cursor of the motion path along the perpendicular orientation of the line of identity of the ellipse, as the vagal activity is elevated. Conversely, the configuration becomes longer and narrower with greater displacement of the cursor of the motion path along the line of identity of the ellipse, as the sympathetic activity is elevated.

The inherent assumption behind using consecutive points in Poincaré plot is that the “present-RR-interval” significantly influences the “following-RR-interval”.

The coupled network oscillator model of cardiac chronotropy describes two distinct dynamic regulatory systems of heart rate (vasomotor-sympathetic and respiratory-vagal) that may be better evaluated by the proposed protocol of dynamic analysis of Poincaré Plot Geometry.

The study of RR interval time series asymmetry is usually done in the Poincaré set up with respect to the line of identity. All points on the line of identity (*x*=*y*) have equal consecutive RR intervals. Hence, any point above the line of identity corresponds to increasing RR interval (i.e. decreasing heart rate) and any point below the line corresponds to decreasing RR interval (i.e. increasing heart rate). Based on this, asymmetry is defined and quantified. This asymmetry can visually be observed by the area of point-clouds above and below the line of identity. In healthy condition, the heart shows continuous variability owing to vagal and sympathetic activities, which impacts on the formation of cloud around the line of identity [36].

However, this definition of asymmetry although encompasses the short-term beat-to-beat variability, it does not represent more sustained increasing or decreasing pattern in the heart rate due to various physiological and other stresses. With more sustained change in heart rate, the Poincaré cloud as a whole, moves downwards or upwards with respect to the line of identity. In the Fig. 4, this becomes clearly visible when the experimental volunteer at rest is subjected to orthostatic stress during a slow head up tilt protocol.

It is reasonable that, scatter points above the line of identity indicate RR intervals that are longer than the preceding RR interval, and points below the line of identity indicate a shorter RR interval than the previous. Accordingly, the dispersion of point perpendicular to the line of identity (the “width of the Poincaré cloud”) reflects the level of beat-to-beat variability. This dispersion can be quantified by the standard deviation of the distances of the points from the line of identity (SD1). Also, the scattered points lying on the length of the line of identity indicate an equal increase and decrease of RR intervals throughout the progress of the whole time series. This range of dispersion of RR interval values is quantified by the standard deviation of points along the line of identity (the “length of the Poincaré cloud”), which is designated as SD2. A distinct advantage of Poincaré plots is their ability to identify beat-to-beat cycles and patterns in data that are difficult to identify with spectral analysis(37, 38).

Khandoker *et al* (2013) analyzed the motion path of the progressively accumulating scatter points with several geometrical shapes and forms and attempted to develop various mathematical descriptors [39].

The proposed descriptor of *motion path analysis* may be using a moving window which tracks the temporal information of the signal. The moving windows may comprise of three or four consecutive points from the Poincaré plot and the area of the triangle or quadrangle formed by these points is computed. This area measures the temporal variation of the points in the window. If the points are aligned on a line then the area is zero, which represents the linear alignment of the points.

The virtual velocity of the moving cursor of the motion path is supposed to provide crucial information regarding the extent of variability of the RR interval time series in short term point-to-point basis. In case of large and irregular spatial separation/variation of the successive points of the Poincaré plot cloud, the virtual velocity of the moving cursor of the motion path will be greater than that of smaller variation of the duplet points of the Poincaré plot cloud.

It is stated earlier that, each point of the Poincaré scatter cloud represents a transition state in a complex dynamical system having a specific combination of sympathetic and parasympathetic attributes of the Autonomic Nervous Tone. Accordingly, the transition from one point to another indicates a shift of this combination status. The detail evaluation of the motion path of the successive scatter points of the Poincaré plot would provide crucial insight in understanding the spatio-temporal dynamics of the Autonomic Nervous System in health and in disease.

We hypothesize that the Motion path analysis of the Poincaré Plot may be utilized for spatio-temporal analysis of RR interval time series that may enlighten us with the sympatho-vagal dynamics and balance providing valuable insight of the autonomic profile.

## Data Availability

All data produced in the present study are available upon reasonable request to the authors

https://youtu.be/tJ9MPZ-u0KI

https://youtu.be/Z8x7S94_9nw

https://youtu.be/1VzFk_Z4WdE

https://youtu.be/ROoqfF7_nwk

## Captions of the supplementary video materials

**Video 1: Progressively accumulating Scatter points in PC Plot**

**Video 2: Motion Path of the Poincare Plot**

**Video 3a: Motion path of Poincare Plot during Rest**

**Video 3b: Motion path of Poincare Plot during Tilt**

## Video links

Video 1: https://youtu.be/tJ9MPZ-u0KI

Video 2: https://youtu.be/Z8x7S94_9nw

Video 3a: https://youtu.be/1VzFk_Z4WdE

Video 3b: https://youtu.be/ROoqfF7_nwk

## Declaration of competing interest

### Declaration

a. The present manuscript is not under consideration elsewhere.
b. None of the manuscript’s contents have been previously published.
c. All authors have read and approved the manuscript and approved its submission to ***Future Cardiology.*** They also agreed the order of authorship.

### Statements relating to our ethics and integrity policies

We pledge to make wise use of our resources and to be good stewards of financial, capital, and human resources. We operate within the letter and spirit of the law and prescribed policies, and strive to avoid impropriety or conflict of interest.

### Data availability statement

We are ready to make available the relevant research data to improve the transparency of research and accelerate the pace of discovery, so that more research can be independently verified or made reproducible, in future.

### Ethics approval statement

The ethical approval for studies involving human subjects was obtained from the Institutional Ethics Committee of the Medical College Kolkata Calcutta, India, vide No. MC/KOL/IEC/NON-SPON/144/11-2015

### Patient consent statement

Written informed consent was obtained from each of the study subjects.

#### Clinical trial registration

NA

#### Permission to reproduce material from other sources

NA

### Plain language summary

The Autonomic Nervous System (ANS) helps maintain the body’s internal balance by regulating functions like heart rate. Heart Rate Variability (HRV) analysis measures small changes between heartbeats to assess ANS activity. One common method, the Poincaré plot, uses a scatter plot to show relationships between consecutive heartbeats. Traditional analysis of this plot focuses on overall shape and statistical measures (SD1 and SD2), but these only capture static features.

This paper introduces a new method called motion path analysis, which tracks how the points on the Poincaré plot move over time. This approach captures both short-term and long-term heart rate changes, providing deeper insight into ANS activity. Tests show that this method detects subtle heart rate shifts under different conditions, like stress. Motion path analysis could improve understanding of heart rate regulation and has potential uses in medical research and diagnostics.

### Article highlight

- Motion path analysis of the Poincaré plot, a new method for assessing heart rate variability (HRV) and autonomic nervous system (ANS) activity.
- Unlike traditional approaches, it tracks heartbeat changes over time.
- Reveals the subtle shifts in autonomic balance.
- The technique improves visualization of heart rate dynamics
- Offers potential applications in clinical diagnostics and ANS research.

